# Genetic spectrum and distinct evolution patterns of SARS-CoV-2

**DOI:** 10.1101/2020.06.16.20132902

**Authors:** Sheng Liu, Jikui Shen, Shuyi Fang, Kailing Li, Juli Liu, Lei Yang, Chang-Deng Hu, Jun Wan

## Abstract

Four signature groups of frequently occurred single-nucleotide variants (SNVs) were identified in over twenty-eight thousand high-quality and high-coverage SARS-CoV-2 complete genome sequences, representing different viral strains. Some SNVs predominated but were mutually exclusively presented in patients from different countries and areas. These major SNV signatures exhibited distinguishable evolution patterns over time. A few hundred patients were detected with multiple viral strain-representing mutations simultaneously, which may stand for possible co-infection or potential homogenous recombination of SARS-CoV-2 in environment or within the viral host. Interestingly nucleotide substitutions among SARS-CoV-2 genomes tended to switch between bat RaTG13 coronavirus sequence and Wuhan-Hu-1 genome, indicating the higher genetic instability or tolerance of mutations on those sites or suggesting that major viral strains might exist between Wuhan-Hu-1 and RaTG13 coronavirus.

## Introduction

A novel betacoronavirus SARS-CoV-2 [1] causing human coronavirus disease 2019 (COVID-19) was first reported in Wuhan, Hubei China in December 2019 [2-4]. The pandemic of SARS-CoV2 has infected more than 12 million people over 180 countries and areas around the world with a death over a half million as of July 9, 2020 [5]. The most vulnerable group in this COVID-19 pandemic is elderly and those with different underlying medical conditions such as malnourished, hypertensive, diabetes, cancer and cardiovascular abnormality [6]. Much effort has been devoted by scientists all over the world to understand the features of SARS-CoV2, particularly the viral genome variations. As was well-known, viral genomic mutations play a key role in propagation of SARS-COV-2 in general. Viral mutation may facilitate to alter the viral infectivity- and pose an additional challenge for detection by the host cell, and thus turns them into important research targets especially in the context of vaccine and drug design. Similar to other viruses, SARS-CoV-2 has been creating random mutations on the genome over time. Only some of mutations were caught and corrected by the virus’s error correction machinery [7]. Analysing these data can potentially monitor the viral transmission routes and identify novel mutations associated with the transmission [8]. For example, Given 103 earlier genome sequence data, at least two clades of SARS-CoV-2 were found to be involved in the global transmission based on T > C mutation on a singleton site at 28144 of the complete genome, which was further termed as S clade (C28144) and L clade (T28144) [9]. Evolutionary analyses suggested S clade appeared to be more related o coronaviruses in animals. Most recently, three major clusters of SNVs involved in the pandemic were found by comparing 160 SARS-CoV-2 genomes [10] with RaTG13 [11]. Researchers also employed standard phylogenomic approaches and compared consensus sequences representing the dominant virus lineage within each infected host [11, 12]. Such information will be of important value for the development of vaccine, transmission monitoring and ultimately the control of the pandemic. However, most of these studies were based on limited numbers of SARS-CoV-2 genomes collected during early pandemic time, which might lead to debating conclusions [10, 13-17]. To date, more than 40,000 SARS-CoV-2 whole genome sequences have been uploaded to the online platform The Global Initiative on Sharing Avian Influenza Data (GISAID) database (https://www.gisaid.org/) [18, 19]. With the availability of increased sample size and longer time of SARS-CoV-2 spreading and developing which has covered almost all countries in the world now, an overview of viral mutation patterns is imperative to provide a comprehensive and updated analysis of the viral genetic variations.

In this study, we took advantage of the mega-datasets collected by GISAID which published almost thirty thousand high-quality SARS-CoV-2 genomes with high coverage until June 15, 2020. Our comprehensive analyses clearly revealed distinct patterns of four major group mutations prominent in different countries and areas, suggesting representative SARS-CoV-2 strains correspondingly. We uncovered novel dynamic transmission and evolution patterns for groups of SARS-CoV-2 variants. A few hundred patients were found to have multiple groups of mutations simultaneously. Comparing with four bat coronavirus genomes, we found that alternations of nucleotides on SARS-CoV-2 genome tend to occur at the same sites where bat coronavirus sequences were different from Wuhan-Hu-1. Strikingly, some nucleotide substitutions on SARS-CoV-2 were apt to be the same as RaTG13 coronavirus sequences. We further investigated protein structure alternations caused by the amino acid (AA) changes due to high-frequent nonsynonymous SNVs. Our novel genome-wide discoveries provided more detailed information and shed the light of studying SARS-CoV-2 which has been clouding over the world.

## Results

### Genetic variants of SARS-CoV-2

We downloaded and analysed 28,212 SARS-CoV-2 complete genome sequences after excluding low-coverage ones from the GISAID database. Using Wuhan-Hu-1 (NCBI Reference Sequence: NC_045512.2, GISAID ID: EPI_ISL_402125) as reference genome, we found that total 12,649 nucleotide sites had single nucleotide variants (SNVs) when compared to reference genome. Majority of SNVs had very low occurrence frequency (Fig 1A), suggesting a high chance of random mutations. Four nucleotide substitutions were detected in over 70% of genome sequences: A23403G, C3037T, C14408T, and C241T. They distributed at distinct SARS-CoV-2 genome locations, on the gene body of Spike, *ORF1a*, and *ORF1ab*, and upstream of *ORF1ab*, respectively. Additionally, there were other 50 unique SNVs arose from larger than 1% of populations (n > 282). It is interesting that some of these frequent SNVs occurred almost simultaneously with concurrent ratio larger than 0.9 (see methods) as shown by blue-line connections in Fig 1A. They may appear across different proteins. For example, A23403G changes an aspartate to a glycine on Spike (D614G) while C14408T converts a proline to a leucine on *ORF1ab* (P4715L). Over 99% of both SNVs were found simultaneously on more than 74% samples, suggesting a biological connection between these concurrent variant sites.

**Fig 1.**
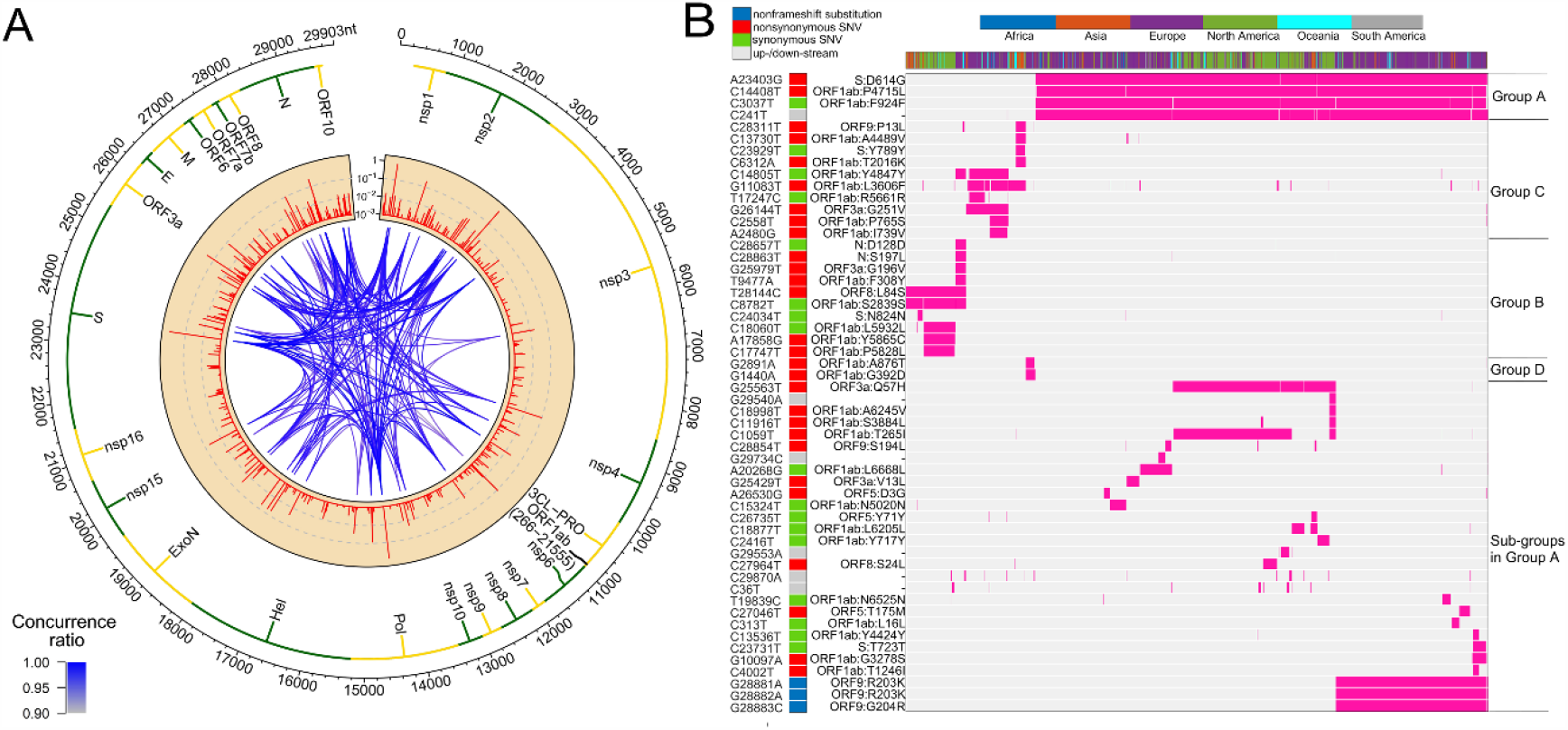
SNVs on about thirty thousand SARS-CoV-2 complete genomes. (A) Circos plot shows distribution, frequency, and co-occurrences of SNV2. From outer to inner circle: coronavirus genome location (nt), gene annotation, occurrence ratios of SNVs at the site (log10 scale, red bars), and connections with high concurrence rates (> 0.9) represented by blue lines. The darker the blue lines, the higher concurrence rates. (B) Fifty-four high frequent SNVs with annotated amino acid (AA) changes were detected (in purple) in about twenty-eight thousand patients worldwide. Four major clusters of SNVs and consequent subgroups can be formed to represent patients from different geographical locations.

Among total 54 frequent SNVs, 31 mutations are nonsynonymous variants or nonframeshift substitutions (Fig 1B). Some of them have been discussed separately by previous studies [9, 11, 20] or marked as elements in clades G, S, and V from the GISAID report [18, 19]. Here, two-way clustering was performed on 54 frequent SNVs and about 28,000 samples (Fig 1B). It is clear to see four major groups of SNVs covering almost all samples, including groups A (C14408T/A23403G, occurring on 21,116 samples), B (T28144C on 2,802 samples), C (G11083T/G26144T on 3,173 samples), and D (G1440A/G2891A on 441 samples). Most SNVs belonged to one unique cluster, while a few SNVs crossed different groups. Taken as an example, a synonymous mutation C14805T existed in both group B and C (Fig 1B), covering over 8% of worldwide samples. Majority (79%) of C14805T can be another signature mutation in group C with SNVs G11083T and G26144T together. In general, the geographical locations of infected patients bearing these special groups of mutations were very different.

Forty countries and areas with numbers of viral genomes larger than 50 were chosen to probe the geographical distributions of these SNVs. Group A, represented by two nonsynonymous mutations, A23403G and/or C14408T, was borne in totally 72% of samples in the study, including about 82% from Europe and 67% from North America (Fig 1B). The top three countries with the highest ratio in group A (Fig 2A) were Russia (99%), Denmark (96%), and South Africa (96%). Group B was distinguished by nonsynonymous mutation T28144C (Fig 1B) which results in substitution of a leucine by a serine on *ORF8*. It was projected in Thailand (50%), Spain (41%), China (31%), and some other Asian countries/areas (Fig 2B). Group C was featured by two nonsynonymous SNVs, G11083T and G26144T (Fig 1B), which substituted a leucine with a phenylalanine on *ORF1ab* and a glycine with a valine on *ORF3a*, respectively. This group existed in many Asian and European countries/areas, e.g. Hong Kong, Singapore, Japan, Turkey etc. (Fig 2C), as reported previously [9, 11, 18-20]. Group D includes two nonsynonymous SNVs, G1440A and G2891A (Fig 1B), both of which change the amino acid sequences on *ORF1ab*. It confirms the clade D, previously defined by Guan et al. [20] based on smaller set of patients. G1440A led to the amino acid change, G212D on nonstructural protein 2 (*nsp2*), while G2891A caused A58T on nonstructural protein 3 (*nsp3*). D-group was mainly found in several European countries, e.g. Wales (17%), Germany (10%), and Belgium (5%) (Fig 2D).

**Fig 2.**
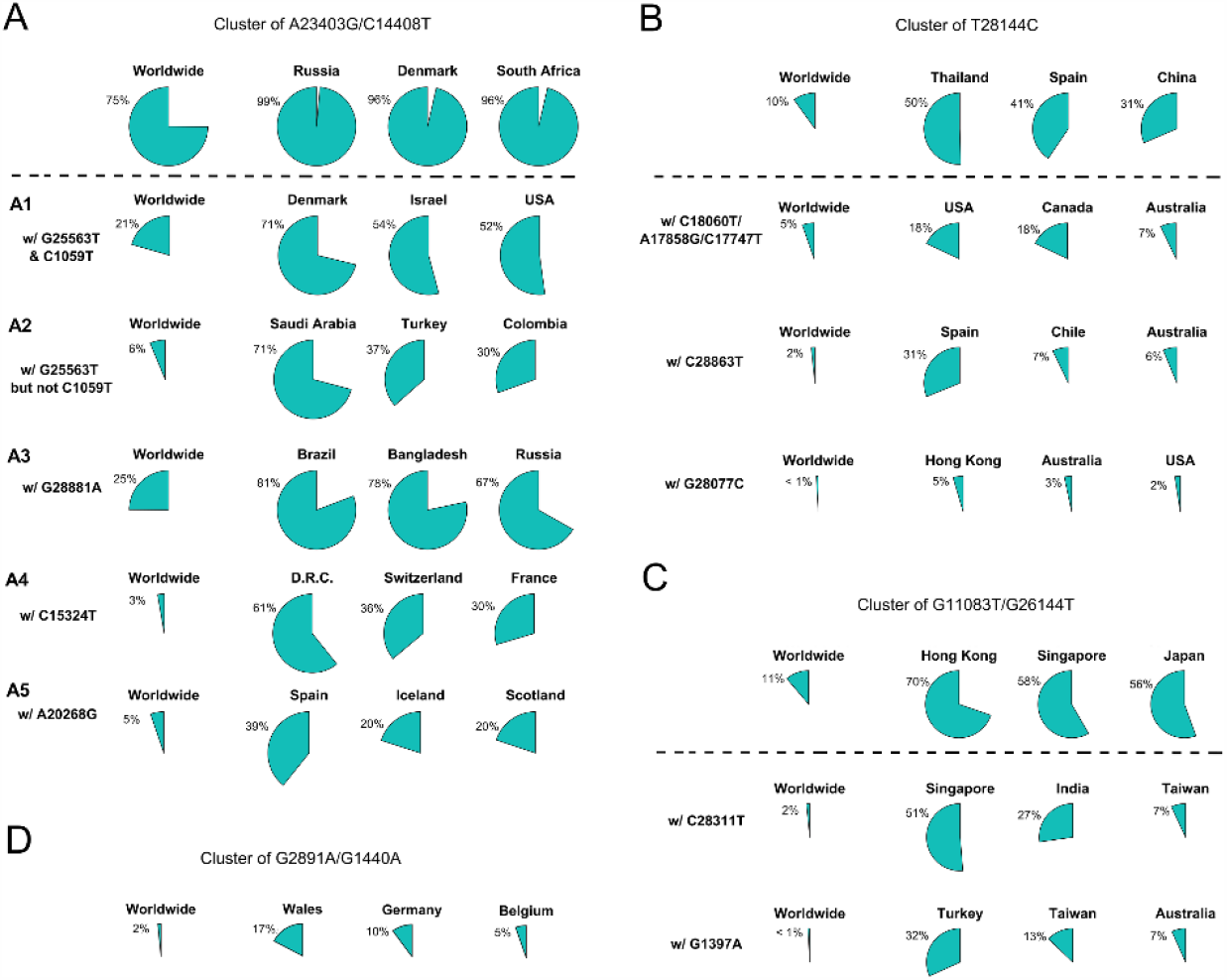
Frequencies of signature SNVs in worldwide and top three countries/areas. (A)-(D) Four major groups and/or their sub-groups had distinct representing countries/areas, indicating different transmission sources and evolution paths.

Besides signature variants in each major group discussed above, some SNVs were found in relatively smaller populations but concurred with the major signature SNVs. Importantly, many non-major SNVs were mutually exclusively presented with each other in different countries and areas (Fig 1B). For instance, about 28 mutations coinciding with A23403G and C14408T in the group A composed sub-types of A (Fig 1B), e.g. G25563T and C1059T. However, two separable sets of samples were associated with different combinations of G25563T and C1059T (Fig 2A). Sub-cluster A1 included both G25563T and C1059T, whereas sub-cluster A2 had G25563T but excluded C1059T. They may represent divergent strains found in distinct populations from varied countries/areas. Specifically, A1 occurred in 21% of all worldwide collected SAR-CoV-2 genomes, particularly in 71% of Denmark, 54% of Israel, and 52% of USA, whereas A2 was found in only 6% population, which were mostly discovered in Saudi Arabia (71%), Turkey (37%), and Columbia (30%). Another sub-cluster, A3, had consecutive mutations at positions 28881-28883 on SARS-CoV-2 complete genome, leading nonframeshift substitutions on *ORF9*: R203K-G204R. A3 occupied 25% of worldwide cases, represented by three major countries, Brazil (81%), Bangladesh (78%), and Russia (67%). Even though some sub-clusters of mutations were found in smaller worldwide populations (around or lesser than 5%), they were significantly over-represented in several countries and areas. For instance, A4 with synonymous mutation C15324T was detected in 61% samples of an African country, Democratic Republic of the Congo (D.R.C.), coming together with 36% of Switzerland and 30% of France.

Patients from one country may have different main groups or sub-types of mutations. A synonymous A20268G in cluster A5 (Fig 2A) was sampled in Spain (39%), Iceland (20%), and Scotland (20%). It is interesting that other 41% of Spain samples had another distinguished nonsynonymous mutation T28144C in group B (Fig 2B), same as many Asian patients had. It suggests the viral transmission path on these patients. 31% of Spain samples also had another unique mutation, C28863T, substituting a serine with a leucine on *ORF9*, concurrent with T28144C. About 18% of Australia samples were found in group B as well (Fig 2B). But they came with additional diverse mutually exclusive SNVs, e.g. either C18060T/A17858G/C17747T (7%), or C28863T (6%), or G28077C (3%). Similar scenarios were observed in USA, where approximately 18% of samples encompassed T28144C with C18060T/A17858G/C17747T, while another 2% was recognized with a different nonsynonymous mutation G28077C in the same main group B (Fig 2B).

SNVs in group C including G11083T and G26144T existed in many Asian countries and areas (Fig 2C), such as Hong Kong, Singapore, Japan, Indian, Taiwan, and Turkey, as reported previously [11]. However, different countries and areas were distinguished by extra variants in the same prime group C. For example, 51% of Singapore was detected with nonsynonymous C28311T on *ORF9*, while Turkey had 32% samples with nonsynonymous G1397A on *ORF1ab*.

Currently, it lacks sufficient evidences to make a conclusive statement about the origins of all SARS-CoV-2 mutations. But time-annotated data collections can still explore geographical evolution patterning of specific SNVs, albeit limited number of high-quality and high-coverage sequenced viral genomes at some time points. For example, only three cases with mutations T28144C and C18060T (one sub-type in group B) reported in Washington State of USA in January 2020, in addition to eight cases in China and additional one in Singapore at almost the same time (Fig 3A). It is notable that T28144C and C18060T concurred with additional de novo nonsynonymous mutations C17747T/A17858G on *ORF1ab* in 52 USA cases in February 2020. No such case was detected in other countries/areas. One month later, this group of signature variants spread over many states of USA, particularly west coast of USA, and other countries and areas of different continents, including Canada, Australia, Iceland, Mexico, New Zealand, and England etc.

**Fig 3.**
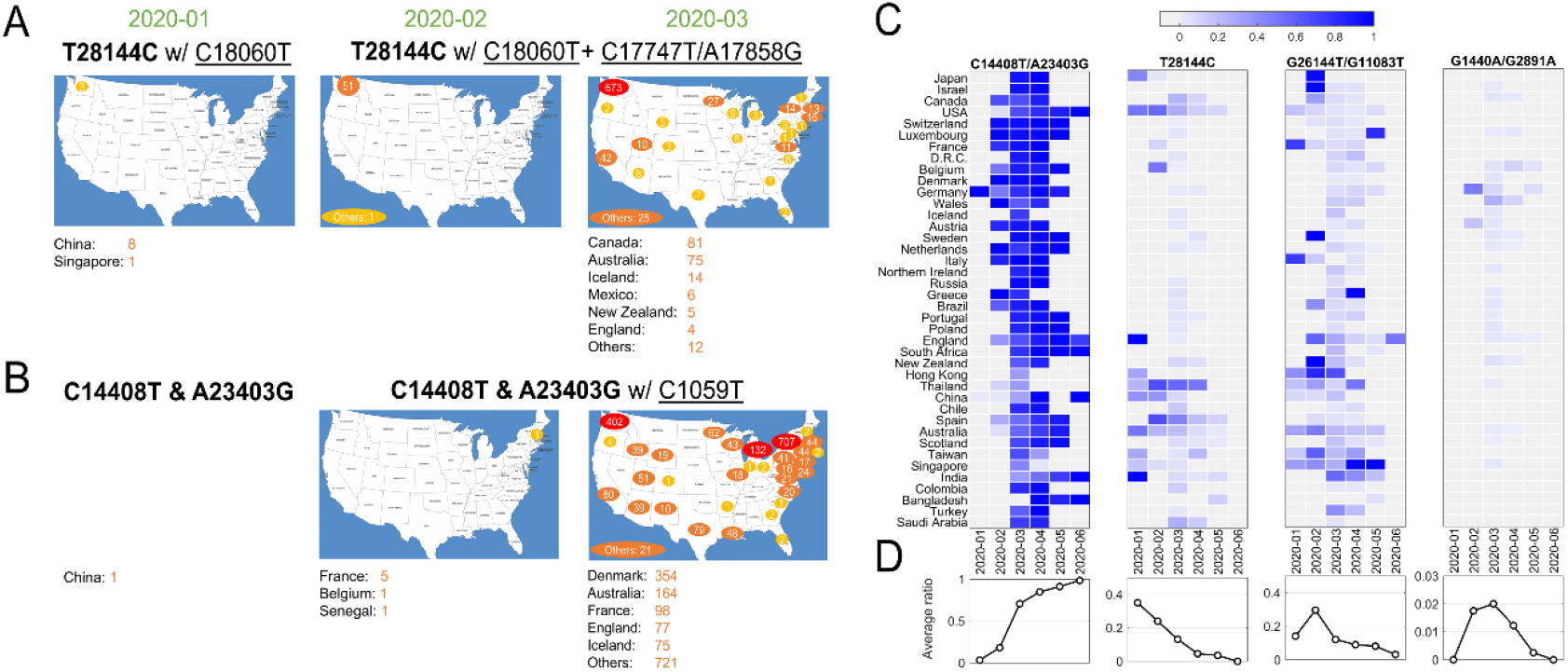
SNVs migration and evolution patterns over time. (A) SNV T28144C with C18060T and additional C17747T/A17858G spread in USA and other countries/areas from January to March of 2020. (B) SNVs of C14408T and A23403G with C1059T spread in USA and other countries/areas from January to March of 2020. (C) The ratios of four significant groups of SNVs, A-D in Figs 1 and 2, varied in different countries/areas with time development. (D) Average temporal ratios of groups A-D SNVs show distinct patterns from January to June of 2020.

Over half of American patients had been sampled with mutations C14408T/A23403G and C1059T on SARS-CoV-2 genome (Fig 2A). Retrieving data in January 2020, we found only one case with both C14408T and A23403G in China from our dataset (Fig 3B). The first case in USA was reported in New Hampshire at the east coast concurrently with C1059T, in addition to five in France, one in Belgium and one in Senegal. The numbers of such cases boosted up in USA and other countries/areas in March 2020, including 354 in Denmark, 164 in Australia, and 98 in France, 77 in England, etc. In the USA, approximately 1,000 cases were found on the east coast of USA, while over 400 cases were identified on the west coast as well.

The variants of group A (C14408T/A23403G) indicated at least two strains of SARS-CoV-2 distinguishable on the sites of Spike and *ORF1ab*. One viral strain observed from Wuhan-Hu-1 can be named as DP with an aspartate on 614 of Spike and a proline on 4715 of *ORF1ab*, while another potential one, named as GL, had a glycine on the site of 614 on Spike and a leucine on the site of 4715 on *ORF1ab* instead. The ratio of GL strain in all USA cases increased dramatically from 6% in February to 87% in May and June 2020 (Fig 3C). The similar growing trend was observed in most of other countries, regardless when this group of mutations were first present (Fig 3C). In general, 91% of samples from all these countries had strain GL since May 2020 compared to only 3% in February (Fig 3D), suggesting that the GL strain of SARS-CoV-2 might become much more stable and prevailing than the other strain DP like Wuhan-Hu-1 after 6-month evolution and transmission.

Different groups of mutations also exhibited distinguished evolution patterns (Figs 3C and D). Taking B-group SNVs for example, we found that the ratio decreased over time from 35% in January 2020 to almost zero in June in these countries, indicating that at least two strains existed at the early of COVID-19 pandemic. However, strains including variant at 28144 other than Wuhan-Hu-1 almost diminished after 7-months of transmission. Only the strain that has the same nucleotide T28144 as Wuhan-Hu-1 finally became the most stabilized strain in the host. The similar patterns were observed for groups C and D as well, even though a sudden increasing was found in February and/or March 2020 due to unknown reasons. For instance, in group C with SNVs G1440A and G2891A, Germany had a high ratio, 47.8% (11 out of 23), in February, while 25% of (96 out of 384) Wales were sampled with the same variants in March 2020.

Four main groups of mutations showed mutually exclusive in about twenty-eight thousand patients, indicating at least five unique viral strains (including the one same as Wuhan-Hu-1) potentially existed in the host. However, as reported in early of March 2020, a patient hospitalized in Iceland infected by two SARS-CoV-2 subtypes simultaneously (https://www.mbs.news/a/2020/03/icelandic-man-reportedly-caught-two-coronavirus-subtypes-simultaneously.html). One strain of the SARS-CoV-2 coronavirus was more aggressive, according to Reykjavik Grapevine newspaper citing CEO of CODE Genetics biopharmaceutical company Kari Stefansson. The second strain is a mutation from the original version of the coronavirus that appeared in Wuhan, China. This was regarded as the first known case of co-infection. Gudbjartsson et. al. [21] reported that patient T25 carries both the A2a1a strain and the A2a1a+25958 strain. As shown in Fig 4, we found that 13 genomes bore both A-group and B-group mutations, while 347 genomes had variant groups of A and C, and both B and C groups were involved in 44 genomes. Strikingly, one patient from Spain was detected with three groups of variants simultaneously, A, B and C. 17 and 4 out of 441 SARS-CoV-2 genomes with D-group SNVs were overlapped with groups A and C, respectively.

**Fig 4.**
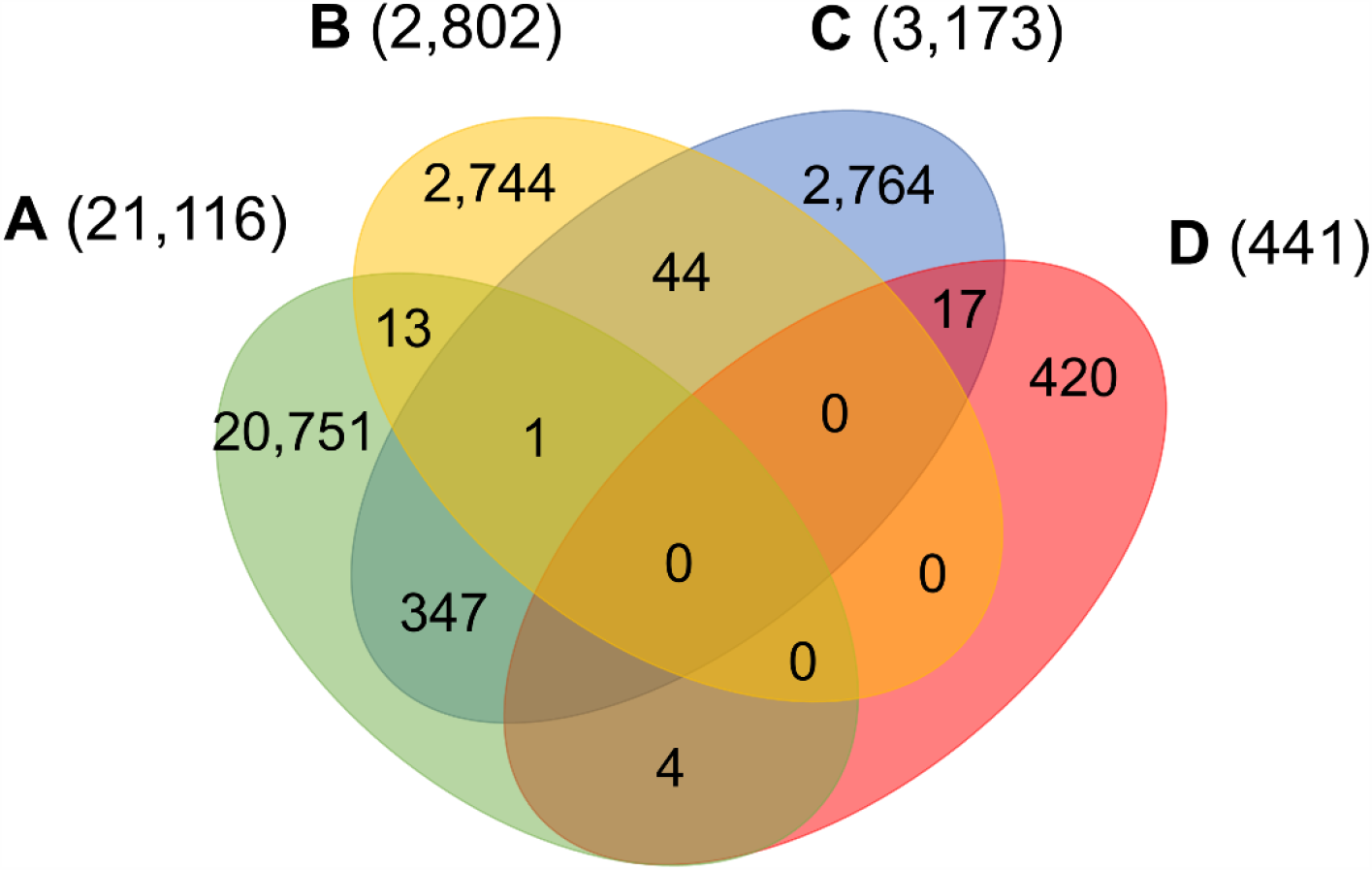
Number of patients carrying significant groups of SNVs, indicating potential co-infection by different SARS-CoV-2 strains.

### Comparison of variants between SARS-CoV-2 genomes and bat coronavirus sequences

Bats were regarded as reservoir species for SARS-CoV-2. To understand potential associations between SNVs among SARS-CoV-2 genomes from patients and bat coronavirus sequences, we also aligned four bat coronavirus sequences to Wuhan-Hu-1 complete genome. The ratios of variants between Wuhan-Hu-1 and bats were 3.8% (RaTG13), 11.1% (bat-SL-CoVZC45), 11.1% (bat-SL-CoVZXC21), and 4.8% (RmYN02). As described above, 12,649 out of 29,903 nts (42.3%) on SARS-CoV-2 genome underwent variation among about 28,000 samples. Interestingly, the ratios of SARS-CoV-2 SNVs on the sites where bats’ sequences differed from Wuhan-Hu-1 were significantly elevated (Fig 5A). Among them, RaTG13 reached the highest ratio (61.5%) with p = 2.7e-40. The result suggests that the sites where Wuhan-Hu-1 differed from bats might have higher tolerance for sequence variations or higher genetic instability.

**Fig 5.**
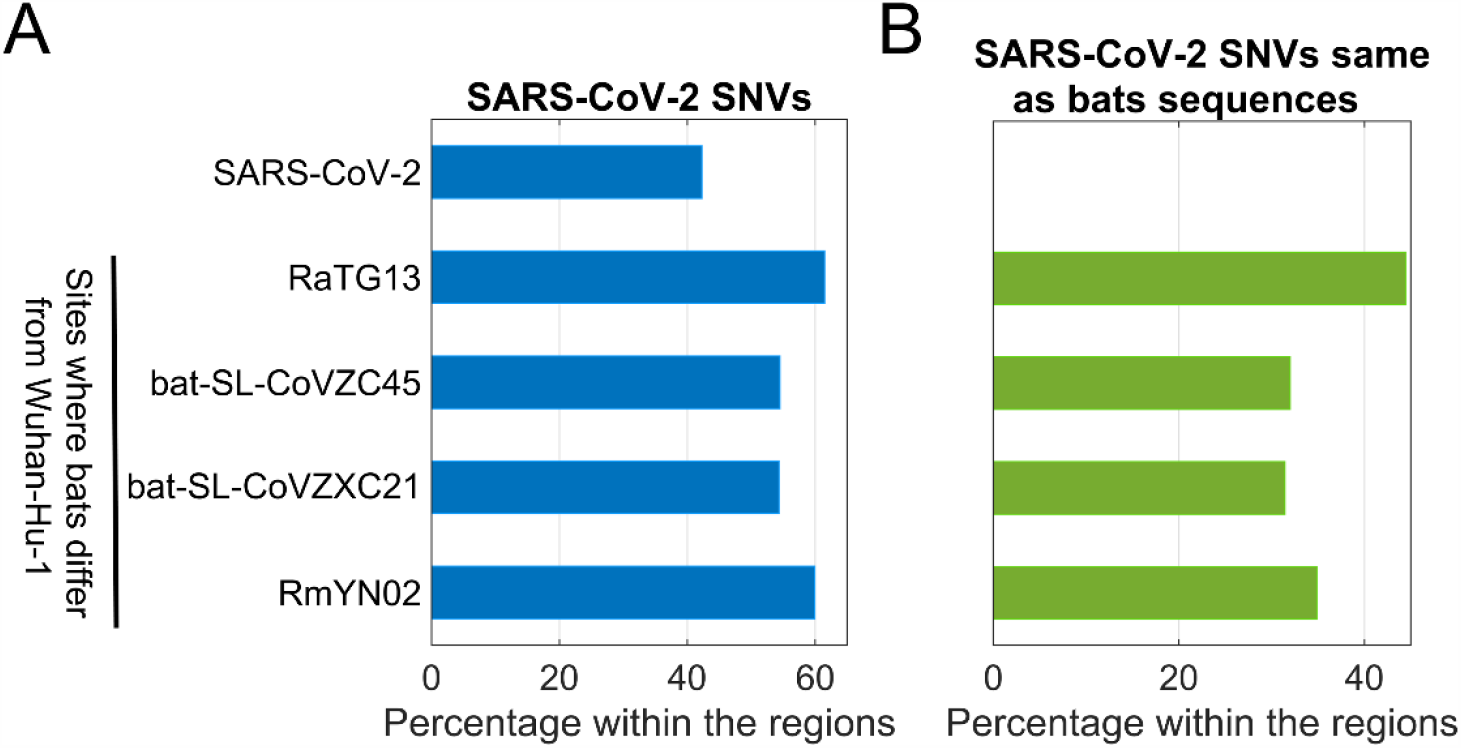
Comparison between SARS-CoV-2 SNVs and nucleotides of four bats coronavirus varying from Wuhan-Hu-1. (A) Percentage of SARS-CoV-2 SNVs on different regions, including SARS-CoV-2 complete genome, and sites where bats coronavirus differ from Wuhan-Hu-1; (B) Percentage of SARS-CoV-2 SNVs which converted to bats nucleotides within the same regions as shown in (A).

In theory, 12,649 identified SARS-CoV-2 SNVs can potentially turn to be any one of three nucleotides other than the original ones from Wuhan-Hu-1. When we focused on the sites where bats coronavirus sequences differed from Wuhan-Hu-1, it turned out that SARS-CoV-2 SNVs had the same mutated nucleotides as RaTG13 coronavirus does on 503 out of 1,132 (44.4%) sites where RaTG13 coronavirus sequence differed from Wuhan-Hu-1 (Fig 5B), including C29095T [11] and seven high frequent SNVs identified from our major groups, e.g. C2416T and C3037T from group A, C8782T, C18060T, C24034T, and T28144C from group B, and C23929T in group C. The ratio for RaTG13 coronavirus was much higher than the ratios observed in other three bat coronavirus sequences (32.0%, 31.4%, and 34.9%, respectively).

### SARS-CoV-2 SNVs and protein functions

Viral sequence mutations will affect the biologic character in the replication, and propagation, and further will alter the toxicity and transmission properties. As previous reported, S68F and P71L non-synonymous mutation in E-protein of SARS CoV-2 were the most common mutation in in E-protein [22]; Q57H, G251V and G196V non-synonymous mutation in *ORF3a* of SARS-CoV-2 would link in the virulence, infectivity, ion channel activity and viral release [23]; deletion of *ORF8* increased the interferon production and reduced in inflammatory cytokines level [24-26]. Most recently, researchers found that D614G non-synonymous mutation located in spike protein would increases infectivity [27-30].

Here, we analyzed the structure change of 31 high-frequent nonsynonymous SNVs and nonframeshift substitutions (Fig 1B) using PyMOL (Schrödinger, Inc.). It was interesting that all of them were on the surface area of corresponding proteins (S1 Table). Eight of them clashed with nearby amino acids on non-structural protein 2 (*nsp2*): G212D, *nsp3*: A58T, *nsp4*: F308Y, *nsp5*: G15S, *ORF3a*: V13L and G251V, *ORF9*: G204R, and *Pol*: A97V, which might be worthy of further analysis. Six of SNVs on *S*: D614G, *ORF3a*: Q57H, *ORF5*: D3G, *ORF9*: G204R, *nsp2*: G212D, *nsp3*: T1198K changed the charge upon the mutations. These changes may contribute to transmission and virulence of SARS-CoV-2. For example, a nonsynonymous SNV, G1440A in group D, discovered in over 400 samples from several European countries, led to the AA change of G212D on *nsp2*. Such a change of G212D may add clashes between residues 212 and ASN183 (Figs 6A and B). It is interesting that *nsp2* G212D falls on the region homologous to the endosome-associated protein similar to the avian infectious bronchitis virus (PDB 3ld1), which plays a key role in the viral pathogenicity [31].

**Fig 6.**
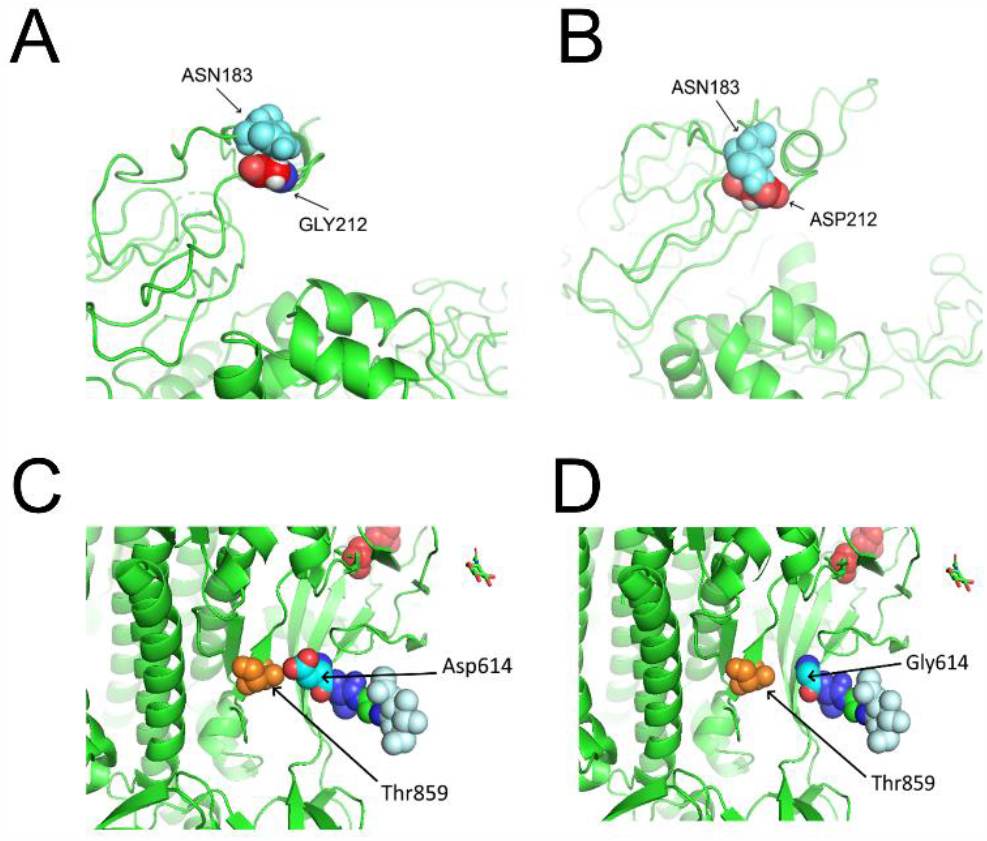
Structures of SARS-CoV-2 non-structural protein 2 (*nsp*2) and Spike near specific mutation sites. (A) No clash was predicted between *nsp2* GLY212 and *nsp2* ASN183 in Wuhan-Hu-1 based on I-Tasser model [57]. (B) A clash was observed between ASP212 and ASN183 after the mutation G212D. (C) There was interaction between T859 and D614. (D) No contact was precited between T859 and G614 after the mutation D614G.

Similar to SARS coronavirus, SARS-CoV-2 entered human cells through its proteolytic proteases and high-affinity receptor-binding domain (RBD) to human protein *ACE2* which enables an efficient cell entry [32]. Furin is responsible for the proteolytic cleavage. In SARS-CoV-2, 15-nt CCTCGGCGGGCACGT encodes five AAs: PRRAR (681-685), locating at 23603-23617 of Wuhan-Hu-1 complete genome. Furin cleavage site bears a RXXR pattern [33, 34]. R685 makes an ideal furin proteolytic cleavage site [35]. Out of almost thirty thousand samples, 58 SARS-CoV-2 genomes were detected with mutations in the region, including 13 from England and 23 from USA. Nonsynonymous SNVs, C23604T, was most frequent among others, causing the mutation of P681L. Other AA mutations included P681H/L/S, R682Q/W, R683P/Q, and A684E/T/S/V. As described previously, D614G on Spike caused by the SNV A23403G in group A covered about three-quarter of total sequenced genomes in our study. D614G on Spike did not generate clashes from the protein structure predictions (Figs 6C and D). However, the residue variations changed the negative polar side chain to neutral nonpolar side chain (Figs 6C and D, S1 Table). Since the site is close to the furin region, such alternation might be able to affect the interactions between furin and furin cleavage sites, then further influence cell-cell fusion and ability to infection [36].

In addition, Cryo-EM-based structural analysis [37] revealed that 5 key amino acids within 434-507 of Spike protein contributed most to the binding activity. This was also confirmed by several recent cryo-EM structural studies [38-41]. The key AAs of SARS-CoV-2 RBD are: L455, G482, V483, E484, G485, F486, Q493, S494 and N501. Interestingly, we identified several nonsynonymous SNVs of SARS-CoV-2 on L455, V483, G485 and S494 from sequenced sample, for instance, G22927T (L455F), G23009T (V483F), T23010C (V483A), G23105A (G485S), and T23042C (S494P). Among them, 28 viral genomes had mutation T23010C (V483A), all of which were sampled in USA, including 26 from Washington State. The RBD for SARS-CoV-2 has residues and motifs found in all three clades in lineage B of betacoronavirus [42], suggesting distinct cell entry pattern than that of other clades. L455, G485, F486, and N501 are among contact points of virus to human *ACE2*, changes in these positions may affect the strength of transmission of the virus.

## Discussion

In this study, we comprehensively analysed almost thirty thousand high-quality and high-coverage SARS-CoV-2 complete genome sequences as well as four bat genome sequences. Even though some SNVs were reported previously and discussed individually, we used bioinformatics approaches to systematically identify four major mutually exclusive groups of SNVs among all samples, suggesting at least five viral strains existing (including one strain same as the reference). These mutations were detected in populations from different geographical locations. The results could provide some insights of possible new functions of SARS-CoV-2 proteins and further bring therapeutic potentials.

Distinct time-course evolution patterns were observed for four major groups of mutations. Some viral strains, e.g. GL with mutations C14408T and A23403G, may gradually replace Wuhan-Hu-1, to become dominant after several month evolutions. Or others may be eliminated naturally with time development, e.g. strains associated with groups B-D mutations (Figs 3C and D). It is hard to explain aberrant emergence of some strains, e. g. the peak time of groups C and D in February and March (Figs 3C and D), particularly due to the lack of enough numbers of high-quality sequenced samples world widely, including China and other countries/areas, before February 2020. However, with more and more clinical data generated, evolution patterning associated with specific biological functions may be clearly uncovered. For example, several groups recently reported that A23403G mutation in Spike protein might alter the antigenic property and transmission ability due to the change of protomer interaction [43, 44].

In general, four SNV clusters were mutually exclusively presented. But we still noticed a few hundred patients who were identified to carry multiple groups of SNVs simultaneously. Without clear evidence that homologous recombination in these regions in the intermediate or human host could occur in these viruses, we just defined such overlaps as co-infections based on our observations and current knowledge. There are several scenarios about these co-infections. One possibility is that two or three strains co-existed and prevailed in the population of the same region during the periods when the patients got infected from other people. Alternatively, the patients could be infected with one strain first then another one later, suggesting that primary infection did not yield immunity in time against the subsequent infection from a different strain. The third possibility is that the virus underwent mutations during the transmission to another human due to the special environment of the host, consequently multiple representative mutations were present on the same patient. Unfortunately, it lacks of information at this moment about the potential post-infection immunity that has important implications for the epidemiologic assessment for the transmission [45]. Of course, the percentage of co-infection cases was less than 1.5% in this study. It might be the consequences of the quarantine and lockdown policy enforced after the spread of COVID-19, while social distancing and wearing face mask are considered effective approaches in reducing the chance of co-infections [46-49]. These policies reduced the likelihood that people met patients with different SARS-CoV-2 strains at the same time.

We further compared SNVs among SARS-CoV-2 genomes from human patients to bat coronavirus sequences. It is interesting that SARS-CoV-2 SNVs, particularly those high-frequent mutations, tend to occur at the same sites where bats coronavirus sequences varied from Wuhan-Hu-1, suggesting the high tolerance of these sites for genetic mutations, or potentials of SARS-CoV-2 turning to a wild-type pathogenic phenotype. RaTG13 coronavirus was most similar to SARS-CoV-2 from perspective of sequences, but it held the highest ratio of SARS-CoV-2 variants which converted to the bat’s coronavirus sequences at the same sites. This suggests that some strains of SARS-CoV-2 deviated from Wuhan-Hu-1 might be more similar to coronavirus in RaTG13 than in other bats presented in this paper. Of course, we don’t have more evidence to show the exact connections between them, but our results may shed the light to search intermediate host and further understand the mechanisms of interspecies transmission in future.

In addition to ORF proteins, four major structure proteins: Spike (*S*), Envelope (*E*), Membrane (*M*) and Nucleocapsid (*N*), help SARS-CoV2 in assembling and releasing new copies of the virus within human cell. We found that all high-frequent SARS-CoV2 SNVs occurred on the surface of proteins. One of most frequent mutations, D614G, has been detected to be dominant around the world now [50]. This SNV caused more infections than other mutations [30]. Korber et al. [44, 50] made suggestions from two frameworks of the potential mechanism of being more infectious: on the structure, D614G disconnects the connection between 614 in S1 and 859 in *S2*, which in turn facilitates the shedding of *S1* from viral-membrane-bound *S2* or impacts RBD-*ACE2* binding by influencing RBD positioning. On the immunological aspect, D614 is within immunogominant linear epitope. Binding of antibody to the epitope may incur conformational change in Spike resulting in nearby RBD enhanced interaction with *ACE2*. Another potential mechanism is due to D614G location. Since D614G is near to furin cleavage sites which are essential for SARS-CoV-2 infection of human, variants on these sites may affect cell-cell fusion and ability to infection [36].

In summary, we attempted to uncover fundamental genetic patterns of SARS-CoV-2 which may help us understand functional consequences due to the viral genetic instability. Our efforts in exploring the views of SARS-CoV-2 migration and evolution in different geographical locations can be helpful to fight against the pandemic. Our findings may provide useful insights on SARS-CoV-2 replication, pathogenicity, and implications. We look forward to incorporating our results with other studies, e.g. interaction maps between SARS-CoV-2 proteins and human proteins [51], for drug discovery, antibody design or vaccine development in near future.

## Materials and Methods

Coronavirus sequences with complete, high coverage, and low coverage excluded were downloaded from GISAID database as of June 15, 2020. 28,212 coronavirus genomes isolated from humans and four bat Rhinolophus affinis were analysed, including Bat CoVRaTG13 and RmYN02 from Yunnan Province, China, SL-CoVZC45, SL-CoVZXC21 from Zhejiang Province, China.. White spaces within the sequences were removed. We aligned these sequences using minimap2 [52] with the reference, the complete genome of Wuhan-Hu-1 (GISAID ID EPI_ISL_402125) by Wu et al. [2, 53]. The variants were annotated by ANNOVAR [54] using NCBI Reference Sequence: NC_045512.2.

Circos plot [55, 56] was made given the ratios of genomes with SNV at each genome location of SARS-CoV-2. The concurrence ratio between two SNVs, X and Y, was defined as the ratio of the numbers of samples with both X and Y to the minimal number of samples with either X or Y.

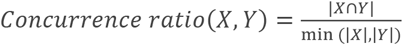

The connection lines in the Circos plot represent SNV pairs with high concurrence ratios (larger than 0.9).

Two-way clustering was performed to categorize the SNVs and samples with a distance function of one minus concurrence ratio on 54 frequent SNVs and about twenty-eight thousand samples.

While comparing SARS-CoV-2 genomic mutation sites and sites where Wuhan-Hu-1 varying from bats’ coronavirus, we used hypergeometric model to calculate the statistical significance of the overlaps.

## Data Availability

Whole genomic sequences of Novel Pneumonia Coronavirus (SARS-CoV-2/hCoV-19/2019-nCoV) were downloaded from the Global Initiative on Sharing All Influenza Data (GISAID) EpiFluTM.

## Acknowledgments

We are grateful to scientists and researchers for depositing whole genomic sequences of Novel Pneumonia Coronavirus (SARS-CoV-2/hCoV-19/2019-nCoV) at the Global Initiative on Sharing All Influenza Data (GISAID) EpiFluTM; Thanks to GISAID database for allowing us to access the sequences for non-commercial scientific research.

---

**S1 Table.** Change of properties of frequent nonsynonymous variants or nonframeshift substitutions.

| Protein | SNV | REF Charge | REF Polar | REF Molar Mass | ALT Charge | ALT Polar | ALT Molar Mass | Side of Surface | Predict Clash | Size change in Molar Mass | Charge change | Polar change | Equivalent protein | Equivalent Protein SNV |
| --- | --- | --- | --- | --- | --- | --- | --- | --- | --- | --- | --- | --- | --- | --- |
| S | D614G | Negative | polar | 132 | neutral | nonpolar | 75 | Outer |  | -57 | Yes | Yes |  |  |
| ORF3a | G251V | neutral | nonpolar | 75 | neutral | nonpolar | 117 | Outer | Leu219 | 42 | No | No |  |  |
| ORF3a | G196V | neutral | nonpolar | 75 | neutral | nonpolar | 117 | Outer |  | 42 | No | No |  |  |
| ORF3a | Q57H | neutral | polar | 146 | Positive | polar | 155 | Outer |  | 9 | Yes | No |  |  |
| ORF3a | V13L | neutral | nonpolar | 117 | neutral | nonpolar | 131 | Outer | Val80 | 14 | No | No |  |  |
| ORF5/M | D3G | Negative | polar | 132 | neutral | nonpolar | 75 | Outer |  | -57 | Yes | Yes |  |  |
| ORF5/M | T175M | neutral | polar | 119 | neutral | nonpolar | 149 | Outer |  | 30 | No | Yes |  |  |
| ORF8 | S24L | neutral | polar | 105 | neutral | nonpolar | 131 | Outer |  | 26 | No | Yes |  |  |
| ORF8 | L84S | neutral | nonpolar | 131 | neutral | polar | 105 | Outer |  | -26 | No | Yes |  |  |
| ORF9/N | P13L | neutral | nonpolar | 115 | neutral | nonpolar | 131 | Outer |  | 16 | No | No |  |  |
| ORF9/N | S194L | neutral | polar | 105 | neutral | nonpolar | 131 | Outer |  | 26 | No | Yes |  |  |
| ORF9/N | S197L | neutral | polar | 105 | neutral | nonpolar | 131 | Outer |  | 26 | No | Yes |  |  |
| ORF9/N | R203K | positive | polar | 175 | positive | polar | 147 | Outer |  | -28 | No | No |  |  |
| ORF9/N | G204R | neutral | nonpolar | 75 | positive | polar | 175 | Inner | Met411 | 100 | Yes | Yes |  |  |
| ORF1ab | T265I | neutral | polar | 119 | neutral | nonpolar | 131 | Outer |  | 12 | No | Yes | Nsp2 | T85I |
| ORF1ab | G392D | neutral | nonpolar | 75 | Negative | polar | 132 | Outer | Asn183 | 57 | Yes | Yes | Nsp2 | G212D |
| ORF1ab | I739V | neutral | nonpolar | 131 | neutral | nonpolar | 117 | Inner |  | -14 | No | No | Nsp2 | I559V |
| ORF1ab | P765S | neutral | nonpolar | 115 | neutral | polar | 105 | Outer |  | -10 | No | Yes | Nsp2 | P585S |
| ORF1ab | A876T | neutral | nonpolar | 89 | neutral | polar | 119 | Outer | Ile62 | 30 | No | Yes | Nsp3 | A58T |
| ORF1ab | T1246I | neutral | polar | 119 | neutral | nonpolar | 131 | Inner |  | 12 | No | Yes | Nsp3 | T428I |
| ORF1ab | T2016K | neutral | polar | 119 | positive | polar | 147 | Outer |  | 28 | Yes | No | Nsp3 | T1198K |
| ORF1ab | F3071Y | neutral | nonpolar | 165 | neutral | polar | 181 | Outer | Phe71 | 16 | No | Yes | Nsp4 | F308Y |
| ORF1ab | G3278S | neutral | nonpolar | 75 | neutral | polar | 105 | Outer | Lys97 | 30 | No | Yes | Nsp5 | G15S |
| ORF1ab | L3606F | neutral | nonpolar | 131 | neutral | nonpolar | 165 | Outer |  | 34 | No | No | Nsp6 | L37F |
| ORF1ab | S3884L | neutral | polar | 105 | neutral | nonpolar | 131 | Outer |  | 26 | No | Yes | Nsp7 | S25L |
| ORF1ab | A4489V | neutral | nonpolar | 89 | neutral | nonpolar | 117 | Outer | Gln117 | 28 | No | No | Pol/nsp12 | A97V |
| ORF1ab | P4715L | neutral | nonpolar | 115 | neutral | nonpolar | 131 | Outer |  | 16 | No | No | Pol/nsp12 | P323L |
| ORF1ab | P5828L | neutral | nonpolar | 115 | neutral | nonpolar | 131 | Outer |  | 16 | No | No | Hel/nsp13 | P504L |
| ORF1ab | Y5865C | neutral | polar | 181 | neutral | polar | 121 | Outer |  | -60 | No | No | Hel/nsp13 | Y541C |
| ORF1ab | A6245V | neutral | nonpolar | 89 | neutral | nonpolar | 117 | Outer |  | 28 | No | No | ExoN/nsp14 | A320V |

## References

1. Viruses, C.S.G.o.t.I.C.o.T.o., The species Severe acute respiratory syndrome-related coronavirus: classifying 2019-nCoV and naming it SARS-CoV-2. Nat Microbiol, 2020. 5(4): p. 536–544.

2. Wu, F., et al., A new coronavirus associated with human respiratory disease in China. Nature, 2020. 579(7798): p. 265–269.

3. Zhou, P., et al., A pneumonia outbreak associated with a new coronavirus of probable bat origin. Nature, 2020. 579(7798): p. 270–273.

4. Zhu, N., et al., A Novel Coronavirus from Patients with Pneumonia in China, 2019. N Engl J Med, 2020. 382(8): p. 727–733.

5. Dong, E., H. Du, and L. Gardner, An interactive web-based dashboard to track COVID-19 in real time. Lancet Infect Dis, 2020. 20(5): p. 533–534.

6. Guzik, T.J., et al., COVID-19 and the cardiovascular system: implications for risk assessment, diagnosis, and treatment options. Cardiovasc Res, 2020.

7. Kupferschmidt, K. and J. Cohen, Race to find COVID-19 treatments accelerates. Science, 2020. 367(6485): p. 1412–1413.

8. Zhang, L., et al., Genomic variations of SARS-CoV-2 suggest multiple outbreak sources of transmission. Medrxiv, 2020.

9. Tang, X., et al., On the origin and continuing evolution of SARS-CoV-2. National Science Review, 2020: p. nwaa036.

10. Sanchez-Pacheco, S.J., et al., Median-joining network analysis of SARS-CoV-2 genomes is neither phylogenetic nor evolutionary. Proc Natl Acad Sci U S A, 2020. 117(23): p. 12518012519.

11. Forster, P., et al., Phylogenetic network analysis of SARS-CoV-2 genomes. Proc Natl Acad Sci U S A, 2020. 117(17): p. 9241–9243.

12. Lai, A., et al., Early phylogenetic estimate of the effective reproduction number of SARS-CoV-2. J Med Virol, 2020.

13. Kupferschmidt, K., https://www.sciencemag.org/news/2020/03/mutations-can-reveal-how-coronavirus-moves-they-re-easy-overinterpret, in Science News. 2020.

14. Forster, P., et al., Reply to Sanchez-Pacheco et al., Chookajorn, and Mavian et al.: Explaining phylogenetic network analysis of SARS-CoV-2 genomes. Proc Natl Acad Sci U S A, 2020. 117(23): p. 12524–12525.

15. Chookajorn, T., Evolving COVID-19 conundrum and its impact. Proc Natl Acad Sci U S A, 2020. 117(23): p. 12520–12521.

16. Rambaut, A., et al., A dynamic nomenclature proposal for SARS-CoV-2 to assist genomic epidemiology. 2020, bioRxiv.

17. Mavian, C., et al., Sampling bias and incorrect rooting make phylogenetic network tracing of SARS-COV-2 infections unreliable.. Proc Natl Acad Sci U S A, 2020. 117(23): p. 12522–12523.

18. Elbe, S. and G. Buckland-Merrett, Data, disease and diplomacy: GISAID’s innovative contribution to global health. Glob Chall, 2017. 1(1): p. 33–46.

19. Shu, Y. and J. McCauley, GISAID: Global initiative on sharing all influenza data - from vision to reality. Euro Surveill, 2017. 22(13).

20. Guan, Q., et al., The genomic variation landscape of globally-circulating clades of SARS-CoV-2 defines a genetic barcoding scheme. biorxiv, 2020.

21. Gudbjartsson, D.F., et al., Spread of SARS-CoV-2 in the Icelandic Population. N Engl J Med, 2020.

22. Hassan, S.S., P.P. Choudhury, and B. Roy, SARS-CoV2 envelope protein: Non-synonymous mutations and its consequences. Genomics, 2020.

23. Issa, E., et al., SARS-CoV-2 and ORF3a: Nonsynonymous Mutations, Functional Domains, and Viral Pathogenesis. mSystems, 2020. 5(3).

24. Gong, Y.N., et al., SARS-CoV-2 genomic surveillance in Taiwan revealed novel ORF8-deletion mutant and clade possibly associated with infections in Middle East. Emerg Microbes Infect, 2020. 9(1): p. 1457–1466.

25. Chen, S., et al., Extended ORF8 Gene Region Is Valuable in the Epidemiological Investigation of Severe Acute Respiratory Syndrome-Similar Coronavirus. J Infect Dis, 2020. 222(2): p. 223–233.

26. Li, J.Y., et al., The ORF6, ORF8 and nucleocapsid proteins of SARS-CoV-2 inhibit type I interferon signaling pathway. Virus Res, 2020. 286: p. 198074.

27. Yurkovetskiy, L., et al., SARS-CoV-2 Spike protein variant D614G increases infectivity and retains sensitivity to antibodies that target the receptor binding domain. bioRxiv, 2020.

28. Zhang, L., et al., The D614G mutation in the SARS-CoV-2 spike protein reduces S1 shedding and increases infectivity. bioRxiv, 2020.

29. Eaaswarkhanth, M., A. Al Madhoun, and F. Al-Mulla, Could the D614G substitution in the SARS-CoV-2 spike (S) protein be associated with higher COVID-19 mortality? Int J Infect Dis, 2020. 96: p. 459–460.

30. Li, Q., et al., The impact of mutations in SARS-CoV-2 spike on viral infectivity and antigenicity. Cell, 2020.

31. Angeletti, S., et al., COVID-2019: The role of the nsp2 and nsp3 in its pathogenesis. Med Virol, 2020.

32. Hoffmann, M., et al., SARS-CoV-2 Cell Entry Depends on ACE2 and TMPRSS2 and Is Blocked by a Clinically Proven Protease Inhibitor. Cell, 2020. 181(2): p. 271-280.e8.

33. Molloy, S.S., et al., Human furin is a calcium-dependent serine endoprotease that recognizes the sequence Arg-X-X-Arg and efficiently cleaves anthrax toxin protective antigen. J Biol Chem, 1992. 267(23): p. 16396–16402.

34. Shiryaev, S.A., et al., High-resolution analysis and functional mapping of cleavage sites and substrate proteins of furin in the human proteomes PLoS One, 2013. 8(1): p. e54290.

35. Coutard, B., et al., The spike glycoprotein of the new coronavirus 2019-nCoV contains a furin-like cleavage site absent in CoV of the same clade. Antiviral Res, 2020. 176: p. 104742.

36. Hoffmann, M., H. Kleine-Weber, and S. Pohlmann, A Multibasic Cleavage Site in the Spike Protein of SARS-CoV-2 Is Essential for Infection of Human Lung Cells. Mol Cell, 2020. 78(4): p. 779–784 e5.

37. Wrapp, D., et al., Cryo-EM structure of the 2019-nCoV spike in the prefusion conformation. Science, 2020. 367(6483): p. 1260–1263.

38. Shang, J., et al., Cell entry mechanisms of SARS-CoV-2. Proc Natl Acad Sci U S A, 2020. 117(21): p. 11727–11734.

39. Wang, Q., et al., Structural and Functional Basis of SARS-CoV-2 Entry by Using Human ACE2. Cell, 2020. 181(4): p. 894-904.e9.

40. Shang, J., et al., Structural basis of receptor recognition by SARS-CoV-2. Nature, 2020. 581(7807): p. 221–224.

41. Yuan, M., et al., A highly conserved cryptic epitope in the receptor binding domains of SARS-CoV-2 and SARS-CoV. Science, 2020. 368(6491): p. 630–633.

42. Letko, M., A. Marzi, and V. Munster, Functional assessment of cell entry and receptor usage for SARS-CoV-2 and other lineage B betacoronaviruses. Nat Microbiol, 2020. 5(4): p. 562–569.

43. Becerra-Flores, M. and T. Cardozo, SARS-CoV-2 viral spike G614 mutation exhibits higher case fatality rate. Int J Clin Pract, 2020: p. e13525.

44. Korber, B., et al., Spike mutation pipeline reveals the emergence of a more transmissible form of SARS-CoV-2. Biorxiv, 2020.

45. Kirkcaldy, R.D., B.A. King, and J.T. Brooks, COVID-19 and Postinfection Immunity: Limited Evidence, Many Remaining Questions. JAMA, 2020.

46. Wang, Y., et al., Reduction of secondary transmission of SARS-CoV-2 in households by face mask use, disinfection and social distancing: a cohort study in Beijing, China. BMJ Glob Health, 2020. 5(5).

47. West, R., et al., Applying principles of behaviour change to reduce SARS-CoV-2 transmission. Nat Hum Behav, 2020. 4(5): p. 451–459.

48. Eikenberry, S.E., et al., To mask or not to mask: Modeling the potential for face mask use by the general public to curtail the COVID-19 pandemic. Infect Dis Model, 2020. 5: p. 293–308.

49. Cheng, V.C., et al., The role of community-wide wearing of face mask for control of coronavirus disease 2019 (COVID-19) epidemic due to SARS-CoV-2. J Infect, 2020.

50. Korber, B., et al., Tracking changes in SARS-CoV-2 Spike: evidence that D614G increases infectivity of the COVID-19 virus. Cell, 2020.

51. Gordon, D.E., et al., A SARS-CoV-2 protein interaction map reveals targets for drug repurposing. Nature, 2020.

52. Li, H., Minimap2: pairwise alignment for nucleotide sequences. Bioinformatics, 2018. 34(18): p. 3094–3100.

53. Wu, F., et al., Author Correction: A new coronavirus associated with human respiratory disease in China. Nature, 2020. 580(7803): p. E7.

54. Wang, K., M. Li, and H. Hakonarson, ANNOVAR: functional annotation of genetic variants from high-throughput sequencing data. Nucleic Acids Res, 2010. 38(16): p. e164.

55. Gu, Z., et al., circlize Implements and enhances circular visualization in R. Bioinformatics, 2014. 30(19): p. 2811–2812.

56. Krzywinski, M., et al., Circos: an information aesthetic for comparative genomics. Genome Res, 2009. 19(9): p. 1639–1645.

57. Yang, J., et al., The I-TASSER Suite: protein structure and function prediction. Nat Methods, 2015. 12(1): p. 7–8.

